# The effect of oxygen on hemodynamic variables in neonates with single ventricle after the Norwood procedure

**DOI:** 10.1101/2025.05.21.25328121

**Authors:** Anders Aronsson, Theodor Skuli Sigurdsson, Lars Lindberg

## Abstract

**Background:** Management of neonates with single ventricle physiology after the Norwood procedure relies on optimizing oxygen delivery and achieving an optimal balance between systemic (Qs) and pulmonary blood flow (Qp). The aim of this study was to determine hemodynamic variables and evaluate their response to different fractions of oxygen (FiO2).

**Methods:** We prospectively determined actively circulating volume (ACV), stroke volume (SV), total cardiac output (CO), shuntfraction (Qp/Qs), and vascular resistances (SVR, PVR) after the Norwood procedure with right ventricle to pulmonary arterial shunt. Measurements were done in 16 neonates by using dilution methodology, where changes in blood velocity due to injection of normal saline were recorded by ultrasound sensors (COstatus, Transonic System Inc). Measurements were performed at stable conditions after sternal closure on sedated and mechanically ventilated neonates at an FiO2 of 0.21, 0.5, and 0.9.

**Results:** The Qp/Qs ratio increased from 1.06 (0.7-1.65) (median, (IQR)) to 1.41 (0.93-1.75) (p <0.001) when FiO2 increased from 0.21 to 0.9. This was solely caused by a decrease in Qs, as Qp remained unchanged. Mean arterial pressure (MAP) and central venous pressure (CVP) were stable and at normal levels at different FiO2. SVR increased and PVR remained unchanged when FiO2 increased from 0.21 to 0.9. The indexed ACV (ACVI) was 50 (45–65) ml/kg.

**Conclusion:** Unresponsiveness of Qp to changes in FiO2 indicates that Qp is regulated by the size of the RV to PA shunt and not by pulmonary vascular vasomotion. A low ACVI and normal blood pressure indicate that these children have a compensated hypovolemia. These findings suggest that increasing the blood volume and reducing the afterload are appropriate treatment strategies rather than pulmonary vasodilation.

**Clinical Perspective:** *What Is New?:*

\# In vivo measurements of hemodynamic parameters after Norwood procedure are possible # Pulmonary blood flow is not amenable for manipulation by increasing oxygen.
\# Patients that have undergone a Norwood procedure have low intravascular volume

*What Are the Clinical Implications?:*

\# Active circulating volume should be optimized
\# Systemic cardiac output will benefit from afterload reduction
\# Oxygen supplementation in the range up to 0.4 fraction of inspired oxygen might be beneficial

## INTRODUCTION

Patients with a functional single-ventricle such as hypoplastic left heart syndrome (HLHS) have to undergo the surgical Norwood procedure. Afterwards there is a postoperative period with risks of serious morbidity and mortality^1,2^. The postoperative period is characterized with hemodynamic instability due to ventricular dysfunction and limited systemic circulation^3,4^.

Postoperative care has therefore, focused on factors to improve ventricular function and balance pulmonary flow (Qp) to systemic flow (Qs)^3,5^. Oxygen can act as a pulmonary vasodilator^6^, effectively dilating the arterioles in the lung and increasing Qp in congenital heart diseases with and without shunt^7^. This response is fully manifested within minutes^8^.

The hemodynamic status also depends on circulating blood volume and total cardiac output (Q), which both can be difficult to evaluate by physical examinations, arterial blood pressure, central venous pressure and/or blood gases^9^. Measurement of oxygen consumption (VO2) and estimation of Qp/Qs ratio by the modified Fick’s equation have been used to guide and ensure a balanced circulation^3,10–12^. This can be misleading since the occurrence of both pulmonary venous desaturation and low systemic venous saturation may result in an erroneous underestimation of the Qp/Qs ratio^5^. Since Qs is essential as a main determinant of successful end-organ perfusion and delivery of oxygen (DO2), underestimation of Qp/Qs can result in inadvertent hypoperfusion.

There is a lack of reliable measurements of hemodynamic variables after the Norwood procedure contributing to an incomplete knowledge of the circulation in the single-ventricle physiology. The small size of the neonates and the precarious postoperative condition makes it difficult to conduct hemodynamic measurements.

A technology that uses ultrasound sensors to detect blood dilution has been developed and validated for assessment of hemodynamics in neonates and children with and without shunt circulation ^13–18^. This technology makes it possible to evaluate ACV, SV, total CO and the Qp/Qs ratio^19,20^. Actively circulating volume are of special interest in these marginal patients since this volume directly impact cardiac preload and cardiac function^21^.

Our aim was to determine these hemodynamic variables and evaluate their response to different fractions of inspired oxygen (FiO2) as it has been demonstrated that increasing FiO2 might influence Qp and affect systemic and pulmonary vascular resistance^7,8,22^. Our hypothesis was that the increase in FiO2 would not affect the hemodynamic variables.

## METHODS

### Patient Demographics

Patients with single-ventricle defects scheduled for a Norwood procedure were eligible for this study. Inclusion criteria were informed parental consent, single ventricle physiology and surgery within the first week after birth. Appropriate preoperative stabilization preceded the surgical procedure. All patients underwent surgical palliation with arch reconstruction, placement of a RV to PA shunt (6 mm), and creating an unobstructed atrial septal communication.

Sixteen neonates were included in this prospective observational study. There were 10 boys and 6 girls. Diagnoses included hypoplastic left heart syndrome (14 patients), double inlet left ventricle (1 patient), congenital corrected transposition with hypoplastic subsystemic chamber (1 patient). Survival rate was 100% at three years follow-up. This study was approved by the Ethics Committee of Lund University, Lund, Sweden (Dnr 2013/636 and Dnr 2016/514). Weight and ages at surgery, sternal closure and measurement in Table 1.

**Table 1.**
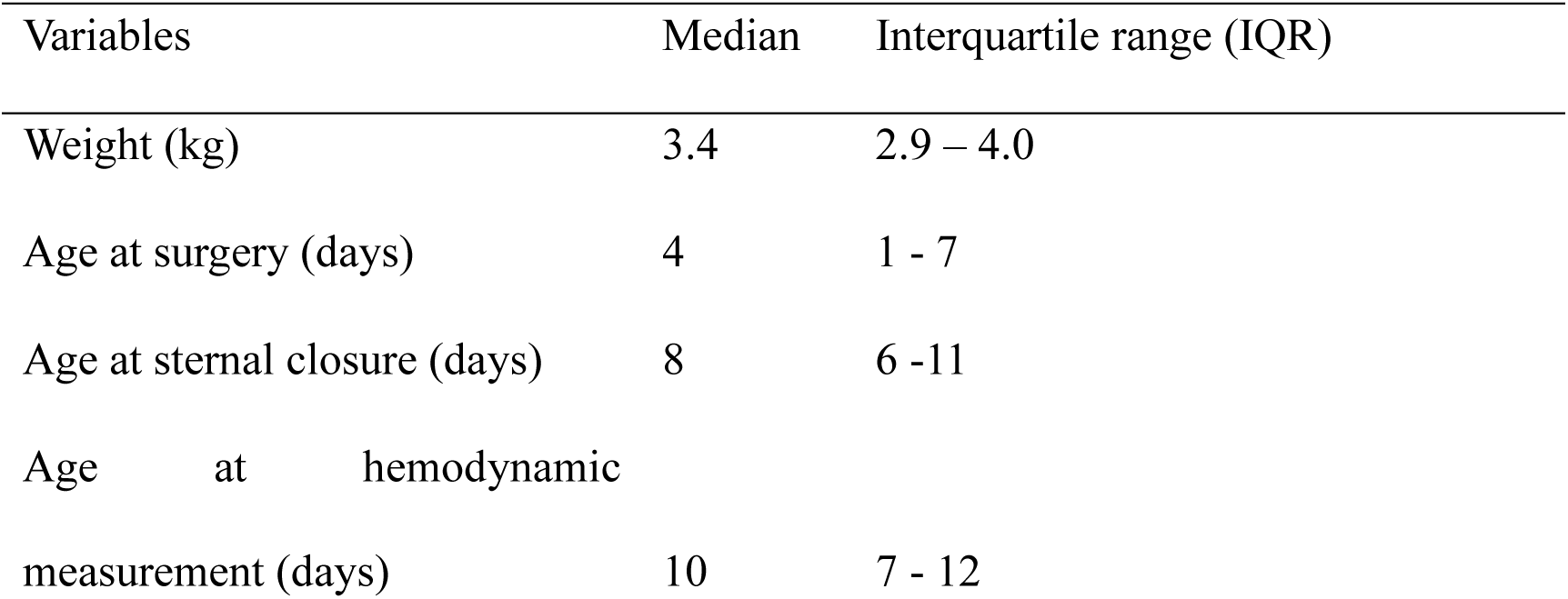
Weight and ages at surgery, sternal closure and measurement.

| Variables | Median | Interquartile range (IQR) |
| --- | --- | --- |
| Weight (kg) | 3.4 | 2.9 – 4.0 |
| Age at surgery (days) | 4 | 1 - 7 |
| Age at sternal closure (days) | 8 | 6 -11 |
| Age at hemodynamic measurement (days) | 10 | 7 - 12 |

### Hemodynamic Measurements

Invasive arterial blood pressure (ABP), CVP, and heart rate (HR) were recorded and arterial and venous blood gases collected. Hemodynamic measurements were made by using saline blood dilution detected by ultrasound sensors (COstatus monitor device,Transonic Systems Inc., Ithaca, NY, USA)^23^. The dilutional curve of the injected saline into the circulatory system is used to calculate the total CO using a modified version of the Stewart-Hamilton equation^14,23^. This is possible since the technology uses an external roller pump which results in a stable blood flow of 10-12 ml/min in an extra-corporeal arteriovenous loop between existing arterial and central venous lines, to which the ultrasound detectors are connected. A rapid rise in the ascending part and/or a delay in the declining part of the dilution curve occur in the single-ventricle due to the mixing of blood between the pulmonary and systemic circulations. The software of the device uses the undistorted part of the dilution curve to create an imaginary plausible area under the curve (AUC) without signs of recirculation representing measured systemic flow (Qs). This AUC can be compared with the actual AUC, representing total CO, which also contains the recirculated blood. This makes it possible to calculate the Qp/Qs ratio ^14^. The method has been validated in small children^13,14^.

Total CO, Qs, and ACV were quantified by the software and Qp/Qs ratio (Qp, Qs), SV, SVR and PVR were calculated. Appropriate hemodynamic variables were indexed to weight (CI, SVI, ACVI, PVRI, and SVRI), since the body surface area (BSA) in children less than 15 kg are non-linear related to body weight^14^.

### Experimental Protocol

Subjects were studied in the pediatric intensive care unit (PICU). During the study they were sedated and mechanically ventilated. All were normothermic (36.4-37.4°C) and had their sternum closed. Sedation was maintained with dexmedetomidine (0.3 – 1.4 mcg/kg/hour), ketobemidone (18 – 30 mcg/kg/hour) and, while the measurements were conducted, propofol (1-3 mg/kg/hour). All neonates were on inotropic support with milrinone (0.5 mcg/kg/minute) and in 4 of the subjects norepinephrine (0.04 – 0.09 mcg/kg/minute). All infusions remained unchanged during the period when measurements were obtained.

Ventilatory support was maintained on ServoI (Maquet) ventilators with pressure regulated volume control (PRVC), tidal volumes of 6-7 ml/kg, and a PEEP of 6-9 cmH2O. Hemodynamic measurements were done at FiO2 0.21, 0.5 and 0.9, starting at FiO2 0.21 and consecutively increased. After inspiratory and expiratory FiO2 had been stabilized according to the oxygen concentration measured by Deltatrac (Datex Ohmeda) and a waiting period of at least five minutes had past, blood gases were drawn and measurements conducted. Blood gas analysis was performed with the ABL800 Flex Radiometer (Radiometer AS, Brönshöj,Denmark). All children had two arterial lines (right radial or ulnar and tibialis posterior arteries) and a 3-lumen central line placed in the right internal jugular vein. Hemodynamic measurements were initiated when circulation was stable and the neonates were immobile. Each measurement session consisted of consecutive repeated measurements with body temperature physiological saline boluses spaced 60 to 120 seconds apart. In one subject movement influenced the stability of the dilution curve, causing rejection of the ACVI. All measurements were obtained by the authors.

### Statistical Analysis

Statistical analysis was performed using SPSS version 29.0.2.0 (IBM SPSS Statistics). No statistical power analysis was conducted before the study as it was designed as convenience sampling.

Shapiro—Wilk test indicates that half of the variables deviate significantly from a normal distribution. Data were therefore expressed as median and interquartile range (IQR). The non-parametric Friedmans analysis of variance (ANOVA) for repeated dependent measurements was used to detect significant differences in the variables between the measurements at 0.21, 0.5 and 0.9. Bon-Ferroni correction was used as the post hoc test to confirm where the differences occurred between the groups.

## RESULTS

### Indexed Actively Circulating Volume

Actively circulating volume index (ACVI) was 50 (45–65) ml/kg, median (IQR) at FiO2 of 0.21. At FiO2 of 0.9 ACVI decreased further to 45 (40-52) ml/kg (p <0.001) (Figure 1).

**Figure 1.**
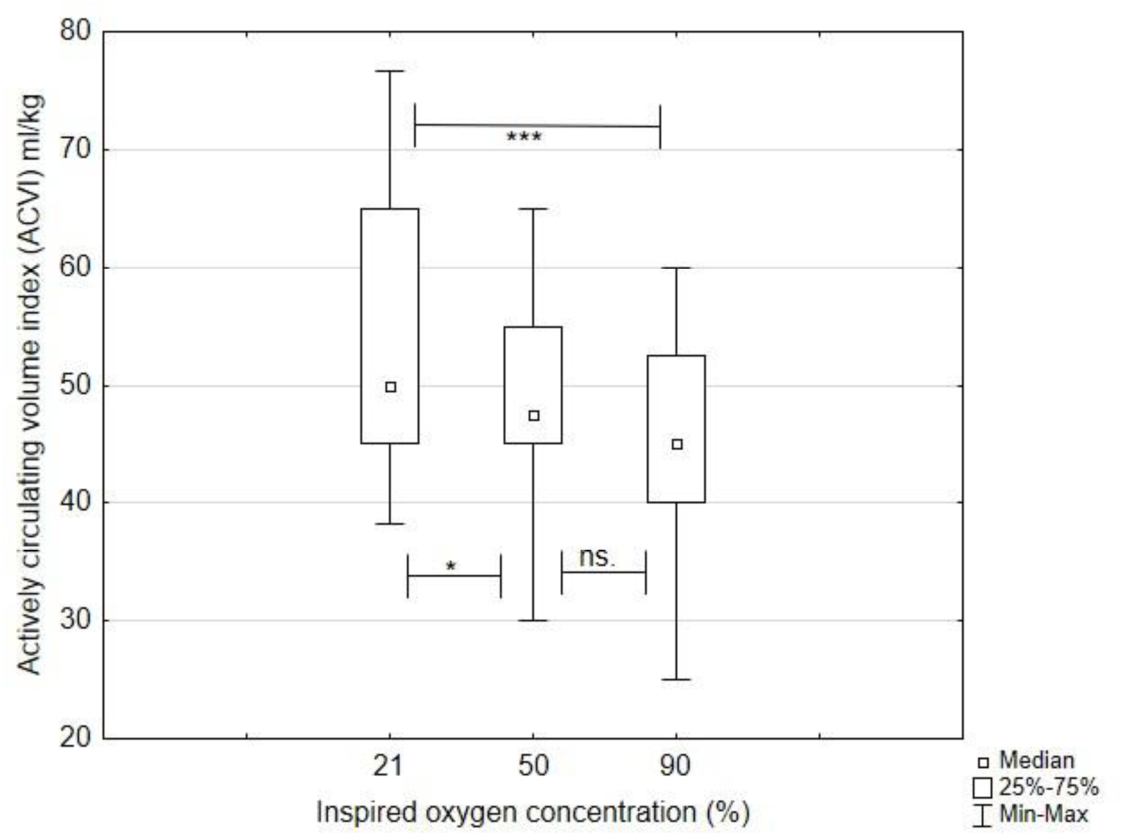
Indexed actively circulating volume (ACVI) at different levels of inspired oxygen concentration (21, 50, and 90 %).

### Indexed Cardiac Output

Cardiac index (CI) was 0.24 (0.22-0.28) L/kg/min at FiO2 of 0.21. CI decreased at higher FiO2 (Figure 2).

**Figure 2.**
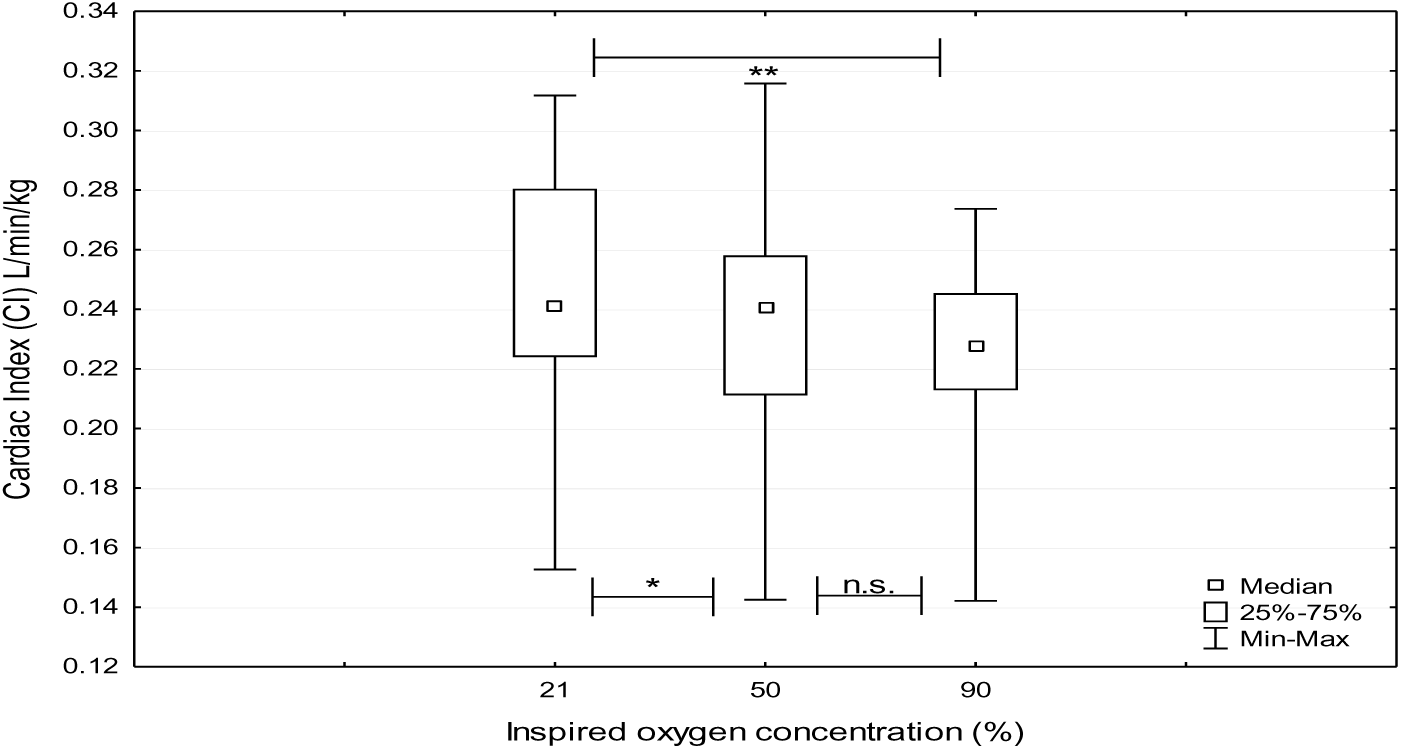
Indexed cardiac output (CI) at different levels of inspired oxygen concentration (21, 50, and 90 %)

Stroke volume index (SVI) was 1.8 (1.6-2.0) ml/kg, and SVI did not change significantly at higher FiO2 levels (Figure 3). The decrease in CI was therefore caused by a decrease in HR at higher FiO2 (Table 2).

**Figure 3.**
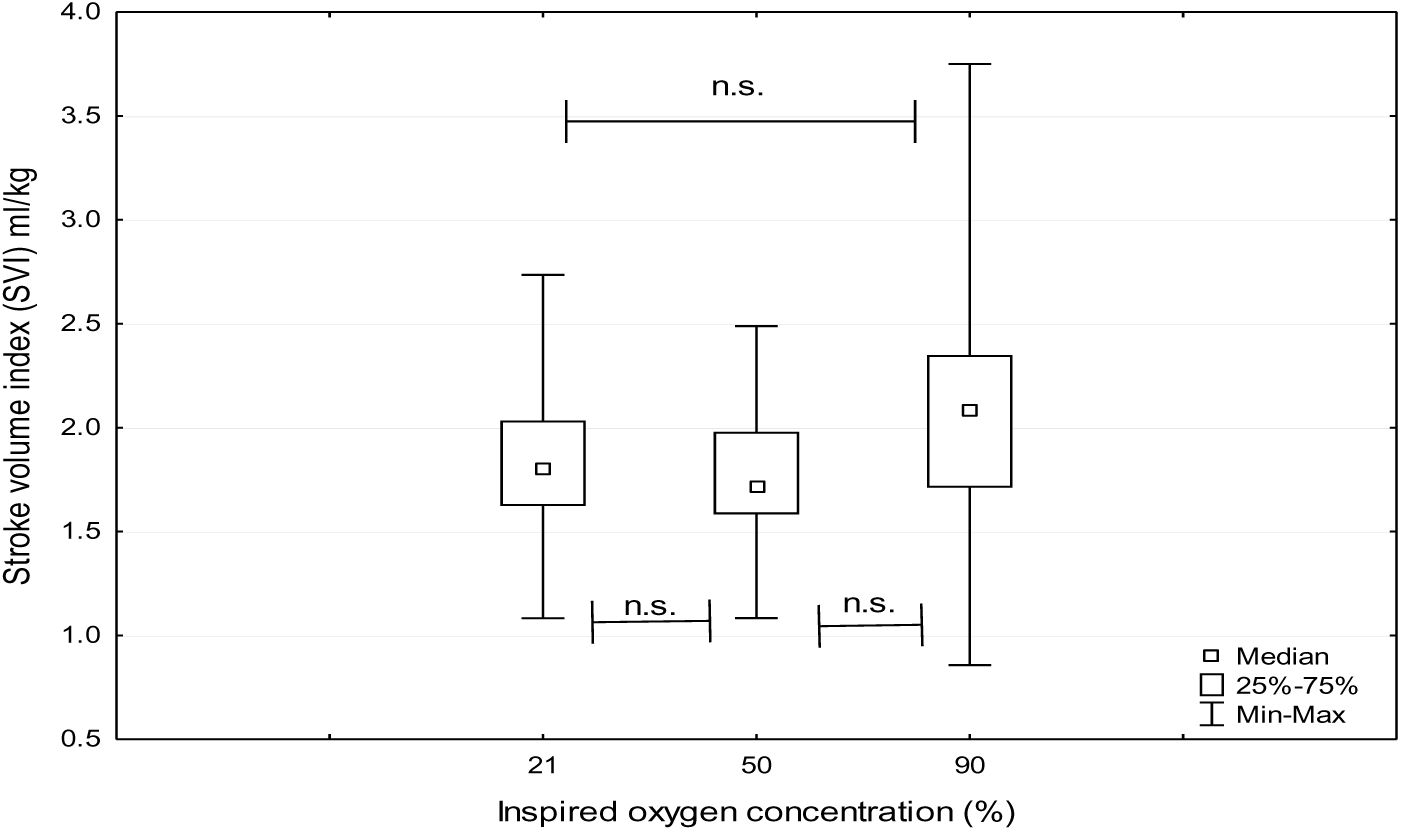
Indexed stroke volume at different levels of inspired oxygen concentration (21, 50, and 90 %)

**Table 2.** Mean arterial pressure (MAP), central venous pressure (CVP), heart rate (HR), hemoglobin, arterial blood saturation (SaO2), venous blood saturation (SvO2) and the arterio-venous saturation difference (SaO2-SvO2) at different levels of inspired oxygen concentration (21, 50, and 90 %). * Friedmans analysis of variance (ANOVA) for repeated dependent measurements

| Variables<br>(median, IQR) | FiO2 0.21 | FiO2 0.5 | FiO2 0.9 | p-value * |
| --- | --- | --- | --- | --- |
| **MAP (mm Hg)** | **47 (45 -51)** | **47 (45 --50)** | **49 (45 - 53)** | **ns.** |
| **CVP (mm Hg)** | **7 (6 – 9)** | **7 (6 – 8)** | **7 (6 – 8)** | **ns.** |
| **HR (beats/min)** | **130 (128 – 146)** | **128 (126 – 144)** | **127 (117 – 141)** | **P < 0.001** |
| **Hemoglobin (g/L)** | **147 (142 – 155)** | **142 (134 - 150)** | **139 (133 - 148)** | **P < 0.001** |
| **SaO2 (%)** | **76 (72 – 79)** | **89 (83 – 90)** | **94 (90 – 96)** | **p < 0.001** |
| **SvO2 (%)** | **55 (42 – 63)** | **69 (58 – 73)** | **73 (62 – 78)** | **p < 0.001** |
| **SaO2-SvO2 (%)** | **18 (15 – 27)** | **20 (17 – 25)** | **23 (16 – 29)** | **ns.** |

Mean arterial pressure (MAP) was 47 (45-51) mm Hg and CVP was 7 (6-9) mm Hg at baseline and did not change significantly throughout the measurements (Table 2).

### Shunt Fraction (Qp/Qs)

Cardiac output (CO) at 21% oxygen was 0.84 (0.76-0.97) L/min. It was divided between the systemic blood flow (Qs), 0.41 (0.29-0.52) L/min (Figure 4) and pulmonary blood flow (Qp), (0.43 (0.30-0.52) L/min (Figure 5) resulting in a Qp/Qs ratio of 1.06 (0.7-1.65) (Figure 6). Qp/Qs increased to 1.41 (0.93-1.75) at FiO2 of 0.9 (p <0.001). This was caused solely by a decrease in Qs, since Qp did not change significantly when FiO2 increased (Figure 4 and 5).

**Figure 4.**
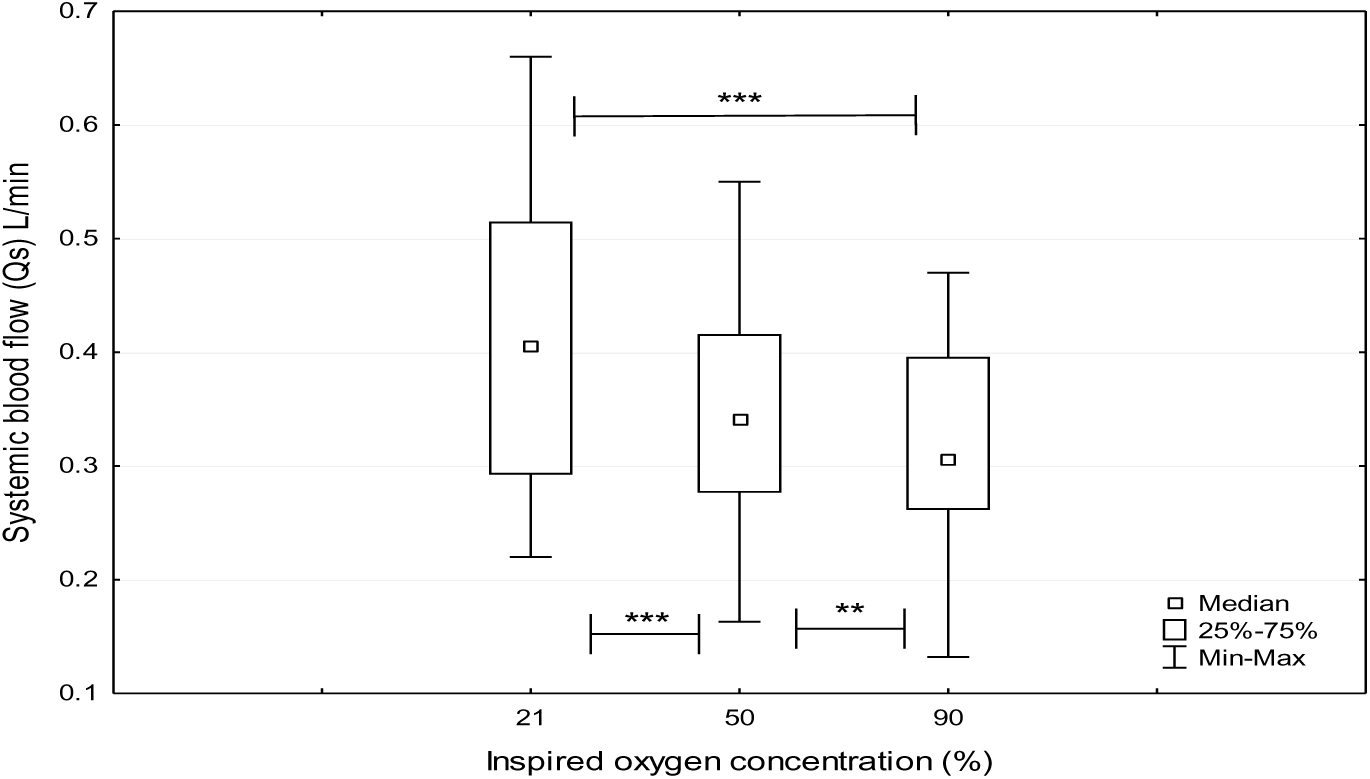
Systemic blood flow (Qs) at different levels of inspired oxygen concentration (21, 50, and 90 %)

**Figure 5.**
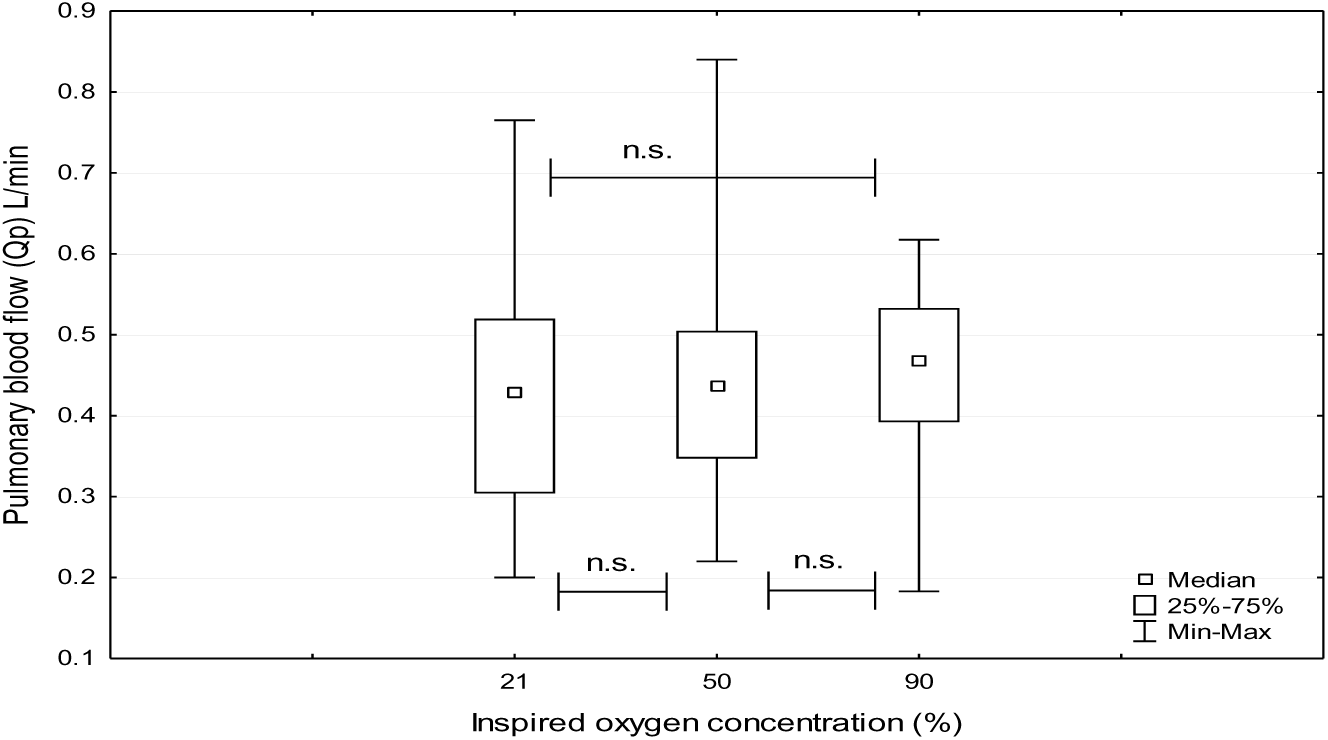
Pulmonary blood flow (Qp) at different levels of inspired oxygen concentration (21, 50, and 90 %)

**Figure 6.**
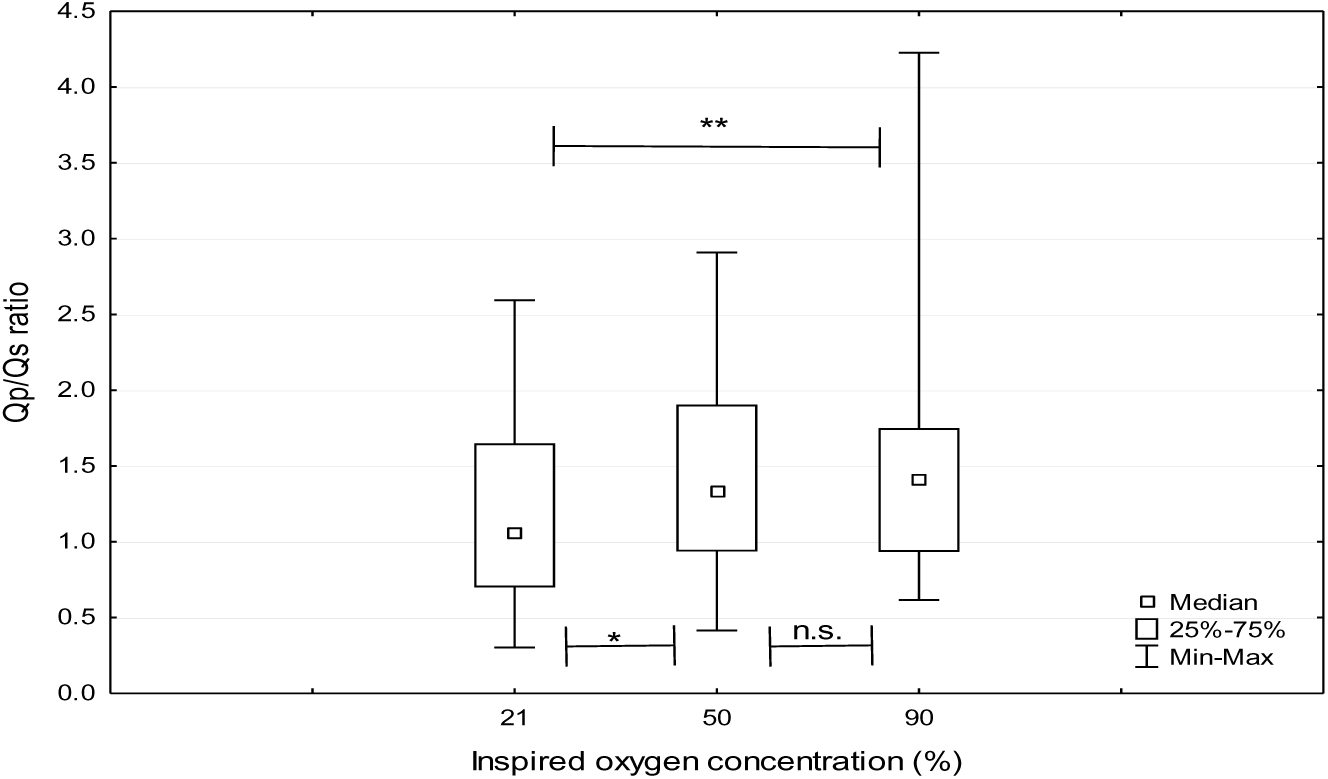
Pulmonary to systemic blood flows (Qp/Qs) at different levels of inspired oxygen concentration (21, 50, and 90 %)

### Vascular Resistances

SVR increased and PVR was unchanged at higher FiO2 (Figure 7 and 8). Since MAP and CVP were unchanged the reason for the increase in SVR was caused by a decrease in measured Qs (Figure 4).

**Figure 7.**
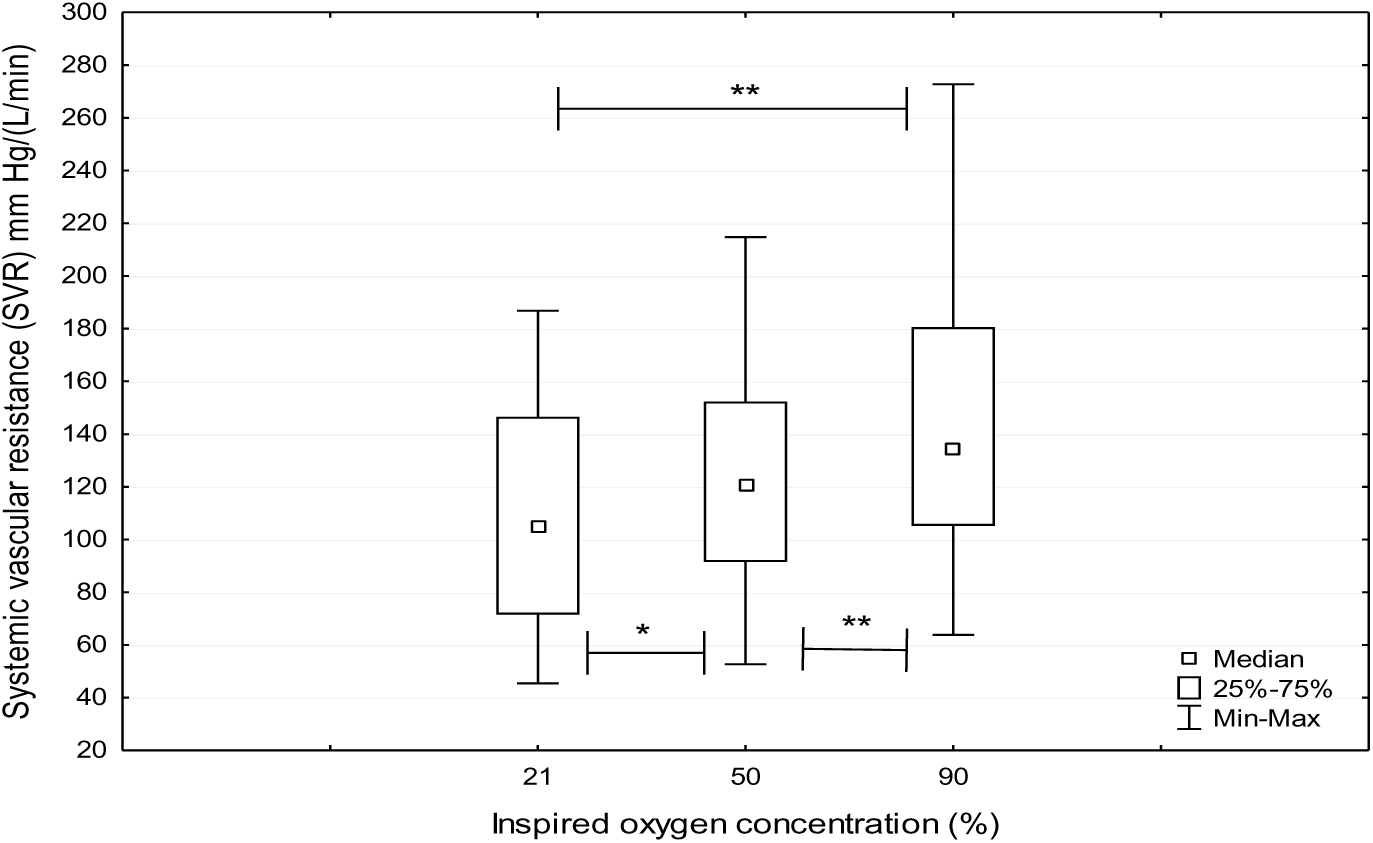
Systemic vascular resistance (SVR) at different levels of inspired oxygen concentration (21, 50, and 90 %).

**Figure 8.**
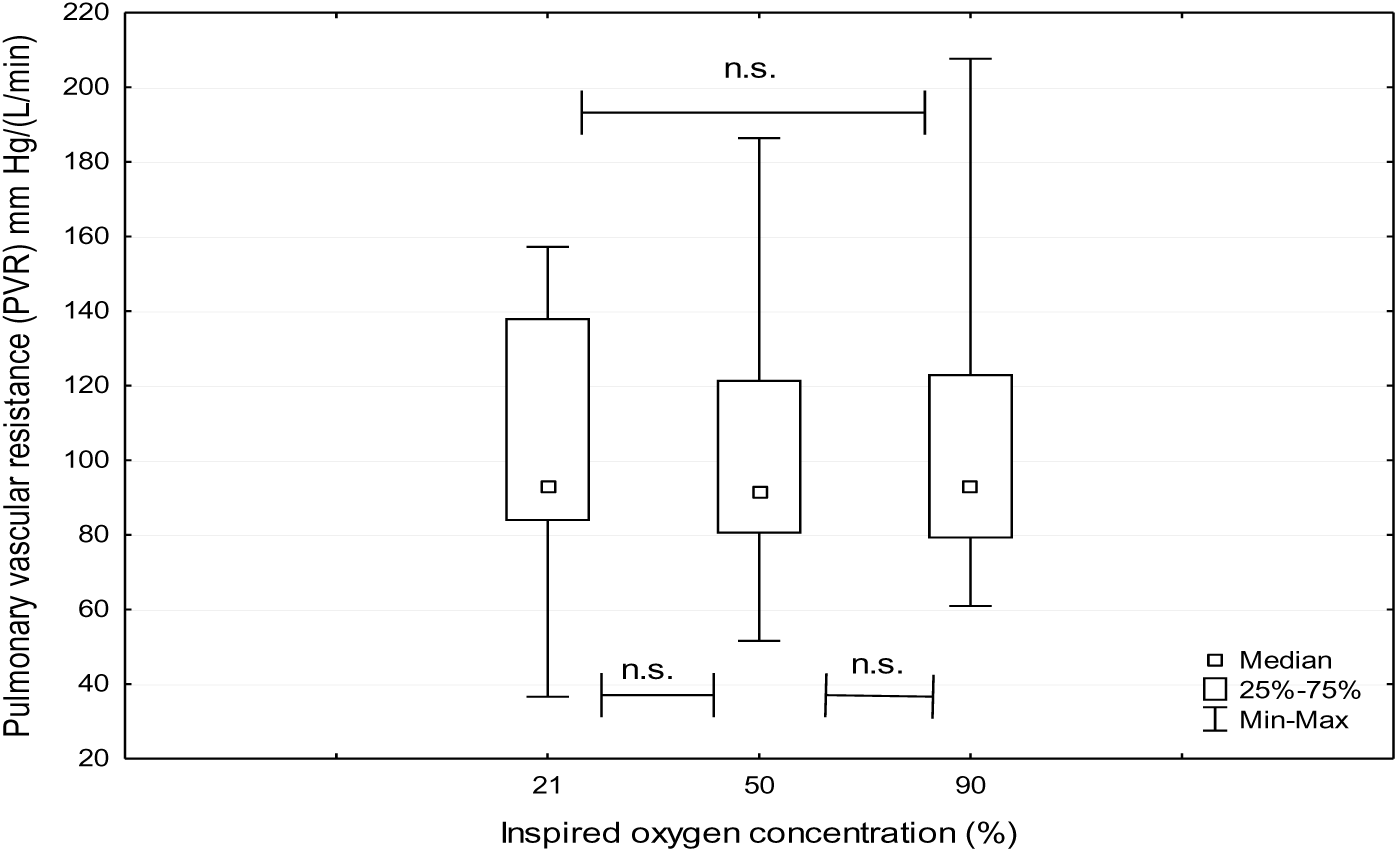
Pulmonary vascular resistance (PVR) at different levels of inspired oxygen concentration (21, 50, and 90 %).

### Oxygen saturations

Arterial oxygen saturation (SaO2) and venous oxygen saturation increased and the arterio-venous saturation difference was unchanged when the oxygen level increased (Table 2).

## DISCUSSION

Our results provide several important insights into the immediate postoperative hemodynamic physiology of the children with single-ventricle after the Norwood procedure with RV to PA shunt. Two observations of interest are evident in this study.

The first observation of interest is that the Qp/Qs ratio increases at higher FiO2, and this was solely caused by a decrease in Qs. Qp was constant at FiO2 between 0.21 to 0.9 indicating that the Qp was dependent on the size of the RV to PA shunt and not influenced by changes in the tonus of the pulmonary vasculature. Notable is that the cardiac index (CI) were 33% higher than in children with biventricular physiology ^14^. Stroke volume index indicates that the single ventricle generated 34 % higher volumes than the left ventricle in children with biventricular physiology ^14^.

Many articles tend to focus on the concept of balancing the pulmonary to systemic circulations in single-ventricle physiology, but our finding indicates that the balancing is a misused concept, since Qp/Qs ratio is only dependent on change in Qs. Increases in inspired level of oxygen did not increase Qp. On the contrary, high levels of oxygen can be detrimental. We found that the driving blood pressure across the pulmonary circulation was stable and pulmonary blood flow was unchanged. If high levels of oxygen dilate the precapillary pulmonary arterioles at a constant pulmonary blood flow without affecting pulmonary arterial blood pressure, it will result in an exposure of the pulmonary capillaries to an increased blood pressure and increased pulmonary capillary filtration pressure. This may increase the leak of fluid into the lung interstitial tissue and increase the diffusion resistance for oxygen across the endothelial-capillary membrane.

In addition, high FiO2 also reduce Qs by systemic vasoconstriction. This vasoconstriction may be unrecognized, since the MAP and CVP were unchanged, but it decreases systemic delivery of oxygen (DO2) at concentrations of FiO2 above 0.5.

Our results agree with the findings of others that neither respiratory alkalosis nor short episodes of increased inspiratory levels of oxygen caused recognizable changes in MAP or CVP in these patient categories ^24,25^. It also agrees with the finding that Qp/Qs was insensitive to manipulation in PVR, and that Qp was limited by the size of the systemic to pulmonary shunt ^26,27^. In addition, a study evaluating the response on the PVR to different levels of PaCO2 found that the PVR was fixed and provided by the systemic to pulmonary shunt ^28^. This may explain why assumed oxygen consumption has been difficult to use as a determinant of cardiac output and results in large errors in the calculated values of PVR ^29,30^. It may also explain the poor correlation between Qp and DO2, SvO2, Sa-vO2, and oxygen excess factor (SaO2/Sa-vO2) ^29^. The second observation of interest is that these children have a significant low ACVI, amounting to 25 % of the ACVI of children scheduled for correction of shunts with biventricular circulation^14^.

This finding occurred despite our policy of a liberal use of blood products and Albumin 5 %. This observation of a low ACVI, which contributed to a systemic hypoperfusion, occurred while the SVR remained high to maintain MAP and CVP. It explains the vulnerability and challenges that may complicate the management of these children. The blood flow from the ventricle to the low pressure pulmonary circulation will be prioritized due to the proximal run-off of the shunt, making Qs extremely blood volume dependent. The slightest decrease in circulatory blood volume, such as can occur during inflammatory responses or infections, may imply that a larger fraction of the existing total blood volume is prioritized to the pulmonary circulation, thereby compromising systemic perfusion and be life-threatening. A low ACV stimulates the renin-angiotensin-aldosterone-system (RAAS) activity to maintain arterial blood pressure by increasing SVR and the sympathetic nervous system activity to maintain preload by increasing venous tonus opposing the need of afterload reduction which has been found to be beneficial^31^.

Children with single ventricle and compensated hypovolemia may be especially vulnerable to afterload reduction with a decrease in diastolic blood pressure and affecting cerebral regional oxygenation^32^. This may be more pronounced in children with lower weight at surgery and those being anemic, who have a lower total blood volume. It is therefore important to balance afterload reduction with an expansion of the circulatory volume in order to compensate for the decrease in SVR.

The postoperative course after Norwood procedure is also characterized by dysoxygenation caused by pulmonary gas exchange abnormalities (ventilation-perfusion mismatch) ^26^. The improvement of SaO2 without change in Qp, when FiO2 increases, indicated that the desaturation is mainly caused by an increased diffusion resistance to oxygen across the alveolar-to-capillary membrane and not due to change in Qp. SaO2 has also been shown to be a poor predictor of Qp/Qs ^26^.

The unresponsiveness to manipulations of the pulmonary vascular tonus also explains why the size of the shunt is of utmost importance to regulate pulmonary blood flow. The surgical decision of the size of the shunt is more important than later attempts trying to regulate pulmonary blood flow by vasodilators^33^. Pulmonary blood flow is limited by the size of the shunt and not by the pulmonary vascular tonus^34^.

Our findings also indicate that SaO2 is mainly dependent on SvO2 which in turn ultimately depends on Qs. Qs is dependent on circulatory blood volume and SVR. This may explain why DO2 is most closely correlated with SVR and Hb and not SaO2 ^12^. These findings therefore affirm postoperative management aimed at maximizing Qs and circulatory blood volume.

It also explains why SaO2 increases temporarily when the children grow and the pulmonary blood flow ratio diverse against Qs rather than Qp during the first months after surgery, before the shunt becomes to restricted and the Glenn anastomosis is needed.

An interesting finding was also the significant increase of SVR at higher FiO2. This has been demonstrated in healthy subjects and in patients with heart failure but not in sedated neonates with single ventricle physiology. A slightly higher FiO2, below 0.4 might be beneficial to overcome the ventilation-perfusion mismatch, but FiO2 at 0.5 and above caused vasoconstriction counteracting afterload reduction and opposed the aim of the management to improve Qs and DO2. Hyperoxia has been demonstrated to have detrimental effects in acyanotic children^35^.

### Limitations

In this study there are several limitations. The number of neonates were limited and they were observed in an early postoperative period when they were sedated and ventilated. The hemodynamic measurements were limited to three levels of FiO2. Hemodynamic data had a wide distribution in this heterogeneous group of study objects which could be expected. Venous blood samples were drawn from superior vena cava and might not reflect a true value of mixed venous saturation.

The shunt ratio estimation was dependent on the mathematical algorithm in the technology used (COstatus). Although, ambiguous dilution curves were rejected, the technology is impaired by a certain inaccuracy and imprecision in the detection of the shunt ratio^18^. The method has been validated in biventricular patients with shunts but not in children with single-ventricle^18^.

## Conclusions

Management in the post-operative period after Norwood procedure should be aimed at increasing Qs by both increasing circulatory volume by blood products or colloids and by an afterload reduction. Based on our data pulmonary vasodilators does not seem to be beneficial since Qp is mechanical regulated by the shunt size.

Our observations indicate that a slight increase in FiO2 can be used without hemodynamic compromises which could be beneficial to overcome the oxygen diffusion resistance across the alveolar-capillary membrane.

## Nonstandard Abbrevations and Acronyms

ABP: Invasive arterial blood pressure
ACVI: Actively circulating volume indexed
AUC: Area under the curve
CI: Cardiac index by weight
CO: Cardiac output
CVP: Central venous pressure
DO2: delivered oxygen
FiO2: Fraction of inspired oxygen
HLHS: Hypoplastic left heart syndrome
HR: Heart rate
MAP: mean arterial pressure
PA: Pulmonary artery
PICU: pediatric intensive care unit
PVR: Pulmonary vascular resistance
Q: cardiac output
Qp: pulmonary blood flow
Qs: systemic blood flow
Qp/Qs: Shunt fraction
RV: Right ventricle
SV: Stroke volume
SVR: Systemic vascular resistance
VO2: Oxygen consumption

## Data Availability

Data available on request

